# Posttraumatic Stress Disorder Epigenome-Wide Association Studies in the Million Veteran Program

**DOI:** 10.64898/2026.09.04.26362177

**Authors:** Sarah Beck, Daniel F. Levey, Marco Galimberti, Cassie Overstreet, Jiarui Chen, Priya Gupta, Cecilia Dao, Bluma J. Lesch, Janitza L. Montalvo-Ortiz, Kelly M. Harrington, The V.A. Million Veteran Program, J. Michael Gaziano, John H. Krystal, Renato Polimanti, Matthew J. Girgenti, Murray B. Stein, Joel Gelernter

**Affiliations:** Department of Psychiatry, VA Connecticut Healthcare System, West Haven, CT, USA; Department of Psychiatry, Yale University School of Medicine, New Haven, CT, USA; Department of Psychiatry, University of California San Diego, La Jolla, CA, USA; Department of Genetics, Yale University School of Medicine, New Haven, CT, USA; Department of Obstetrics, Gynecology, and Reproductive Sciences, Yale University School of Medicine, New Haven, CT, USA; Million Veteran Program (MVP) Coordinating Center, Veterans Affairs Healthcare System, Boston, MA, USA; Department of Psychiatry, Boston University Chobanian and Avedisian School of Medicine, Boston, MA, USA; Division of Aging, Brigham and Women’s Hospital and Harvard Medical School, Boston, MA, USA; Department of Medicine, Harvard Medical School, Boston, MA, USA; School of Public Health, University of California San Diego, La Jolla, CA, USA; VA San Diego Healthcare System, San Diego, CA, USA; Department of Neuroscience, Yale University School of Medicine, New Haven, CT, USA

## Abstract

Genomic studies have improved our understanding of posttraumatic stress disorder (PTSD) biology. Epigenetic differences in DNA methylation can reflect environmental influences, which are critical in PTSD etiology, and may additionally differentiate between PTSD cases and controls or distinguish severity of PTSD symptoms. We conducted epigenome-wide association studies (EWAS) of PTSD in 22,141 subjects from the United States V.A. Million Veteran Program (MVP), using the PTSD Checklist (PCL-C), including 17,674 European ancestry (EUR), 3,430 African ancestry (AFR), and 1,037 admixed American descent (AMR) individuals. We evaluated the 17-item PCL-C and its symptom subdomains (re-experiencing, avoidance, and hyperarousal) and used our previously-reported algorithmically-defined PTSD diagnoses validated against clinical records. There were 9,413 total cases of Lifetime PTSD and 27,257 controls, with 5,184 cases and 19,382 controls in EUR, 3,095 cases and 6,240 controls in AFR, and 1,134 cases and 1,635 controls in AMR. We identified a total of 272 epigenome-wide-significant associated CpG sites across all PTSD traits, or 28 after adjusting for smoking status. We replicated 5 out of 11 CpG sites reported in a prior (Psychiatric Genomics Consortium) PTSD EWAS. There was significant enrichment of genes downregulated in somatostatin interneurons and of genes expressed in GABAergic interneurons. Regression analyses showed that PTSD case status was significantly correlated with accelerated DNA methylation aging. Contrary to previous PTSD EWAS findings, smoking status stratification suggested that the association between PTSD and methylation of the *AHRR* gene may be confounded by smoking status. This is the largest-scale PTSD EWAS in multiple ancestries and the first large PTSD EWAS on symptom severity. We replicate findings from previous EWAS and greatly extend knowledge of the epigenetics of PTSD.

## Introduction

Posttraumatic stress disorder (PTSD) can occur after experiencing or witnessing a traumatic event and, per DSM-IV diagnostic criteria used in the Million Veteran Program (MVP) sample, causes symptoms of re-experiencing (REX), avoidance (AVOID), and hyperarousal (HYP).^1^ It is associated with lower quality of life, higher likelihood of substance use and suicidality, and higher risk for numerous other psychiatric and medical disorders.^2,3^ Approximately 70% of people experience serious trauma in their lifetimes, but only approximately 5% develop PTSD^4,5^, suggesting that there are factors that modify risk. Previous genome-wide association studies (GWAS) have identified numerous genetic risk variants for PTSD and its symptom subdomains.^6-8^

Trauma exposure is integral to PTSD diagnosis. The environmental factors that influence PTSD risk may act via mechanisms that affect gene regulation. The best-studied of these is DNA methylation at cytosine-guanine dinucleotides (CpG sites), which can modulate gene expression and is stably maintained through successive cell divisions.^9^ Maintenance of DNA methylation in regulatory regions of the genome means that epigenetic information can persist long after the initial exposure, providing a molecular record of past biological or environmental influences.

Exposure to trauma has been shown to affect epigenetic marks.^10^ There have been several prior epigenome-wide association studies (EWAS) and meta-EWAS of PTSD^11-13^, the largest of which included 5,077 subjects of European (EUR) and African (AFR) ancestry from 23 studies and identified significant CpG sites in genes related to the immune system and neurotransmission. The most recent EWAS meta-analyses included 2,156 PTSD cases and 2,921 controls from 23 civilian and military studies.^11^ They identified 11 CpG sites associated with PTSD, nine of which were novel, and replicated two findings of a prior epigenome-wide meta-analysis^13^ that included 10 civilian and military studies and reported lower *AHRR* methylation in individuals with PTSD. *AHRR* methylation is a notable characteristic of smoking traits^14^ and can be a more reliable indicator of smoking than self-report.^15^ Accordingly, it is very hard to adjust EWAS for smoking exposure to account for *AHRR* signals. Previous studies that reported PTSD associations with this locus may not have adequately controlled for smoking.

Sample size and heterogeneity were also limitations in prior studies. Here, we address these limitations by conducting the largest EWAS of PTSD to date within a single well-characterized cohort, thereby increasing statistical power and reducing cross-study heterogeneity. The MVP is comprised of military veterans, includes many individuals with PTSD, and has been a major contributor to prior GWAS.^6,7^ The MVP includes several kinds of PTSD-relevant data, including electronic health record (EHR) diagnoses and the self-reported PTSD Checklist for DSM-IV, Civilian version (PCL-C, sometimes designated PCL-17).^16^ We previously reported algorithmically defined case-control status.^17^ The PCL-C allowed us to evaluate PTSD as a quantitative trait and investigate DSM-IV symptom subdomains, as we did previously in a GWAS.^7^ Our study also extends epigenetic analyses of PTSD to larger, more informative samples of non-EUR participants; we include data from EUR, AFR, and AMR. Finally, we quantified methylation-based biological aging using a discrepancy-based approach, regressing Horvath, Levine, and DunedinPACE DNA methylation age or rate of aging onto chronological age to derive age acceleration residuals and evaluate their correlation with PTSD case status.

## Results

### PTSD epigenome-wide associations

In the primary analyses, we identified the following epigenome-wide significant sites for EUR: 148 CpG sites associated with total PCL score (PCL-Total; Figure 1), 29 associated with REX, 263 associated with AVOID (Supplementary Figure 1), 64 associated with HYP, and 4 associated with lifetime PTSD diagnosis (PTSD-Dx), for a total of 269 unique CpG sites significant in EUR (Supplementary Table 1). Significance was defined as a BACON- and Bonferroni-corrected *p*-value < 6.51×10^−8^ (0.05/768,569 tests).^18^

**Figure 1.**
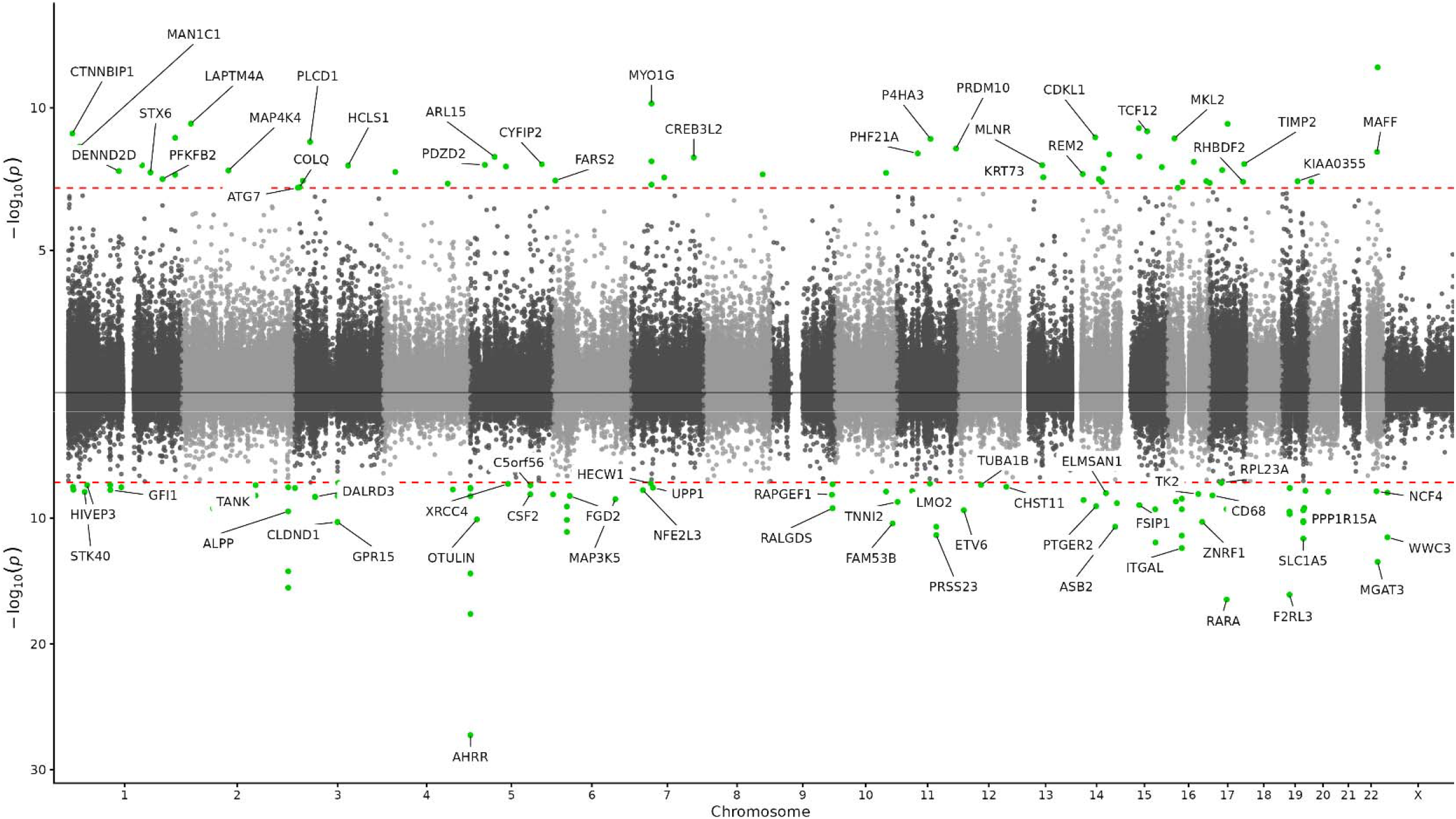
Miami plot for PCL-Total in EUR (N = 17,674) with positive associations on top and negative on the bottom. Statistics were adjusted for inflation and bias using BACON.18

Because hypomethylation of *AHRR* CpG sites is associated with cigarette smoking—this is one of the best-replicated findings in epigenetics—we performed sensitivity analyses to test for the influence of smoking status on our results.^19^ When we adjusted for smoking status, 11 CpG sites were associated with PCL-Total (Figure 2), all of which were significant in the primary analysis except for cg25889918 (*CCDC141*, coefficient with smoking adjustment = 1.31×10^−4^, *p*-value with smoking adjustment = 4.81×10^−8^), which was, however, significant in the primary analysis of AVOID (coefficient = 3.00×10^−4^, *p*-value = 2.43×10^−8^). Twenty-seven CpG sites were associated with AVOID (Supplementary Figure 2), all of which were significant in the primary analysis except cg00994936 (*DAZAP1*, coefficient with smoking adjustment = 3.26×10^−4^, *p*-value with smoking adjustment = 1.65×10^−8^). Two CpG sites were associated with HYP when adjusted for smoking status, but they were captured in other analyses. No CpG sites were associated with REX or PTSD-Dx when we adjusted for smoking status. There was a total of 27 unique CpG sites significant in EUR after adjustment for smoking status (Supplementary Table 2).

**Figure 2.**
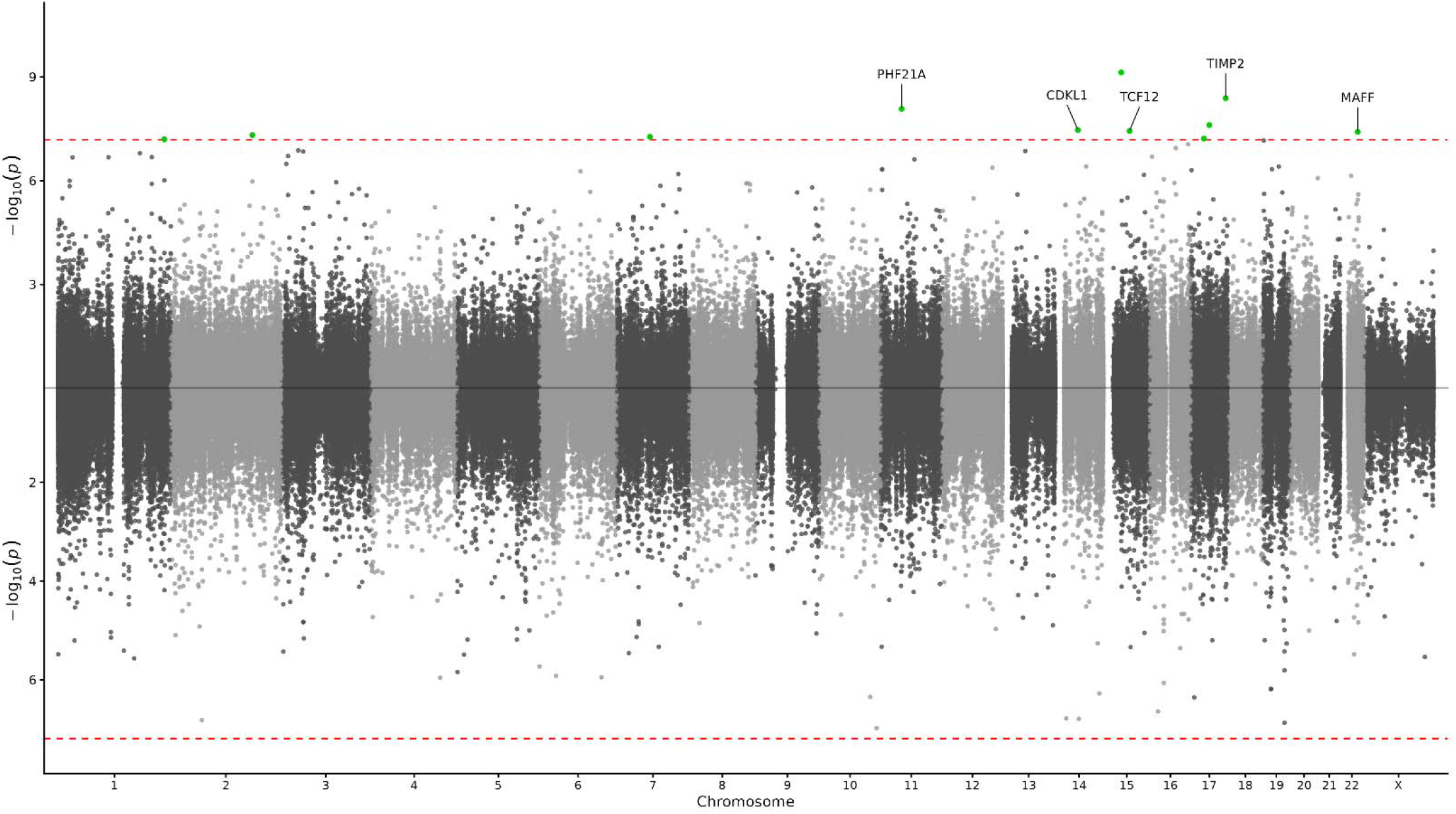
Miami plot for PCL-Total in EUR with smoking status as a covariate (N = 16,542). Positive associations are on top and negative on the bottom. Statistics were adjusted for inflation and bias using BACON.^18^

In AFR, there were no significant CpG sites for PCL-Total, HYP, AVOID, or REX in the primary or the smoking-adjusted analyses. In the primary analysis, PTSD-Dx had two significant associations—cg25322382 (*DDI2*, coefficient = −1.22×10^−3^, *p*-value = 2.83×10^−8^) and cg10213873 (coefficient = −3.78×10^−3^, *p*-value 1.17×10^−8^)—but they were not significant after smoking adjustment. In AMR, REX was associated with cg22036458 in the primary analysis (coefficient = 8.84×10^−4^, *p*-value = 8.19×10^−9^), and this survived smoking adjustment (coefficient = 8.61×10^−4^, *p*-value = 2.07×10^−8^).

Across all ancestries, in the primary analyses, we identified 272 unique CpG sites and 217 genes associated with PTSD. In almost all cases, genomic inflation factors were lower after smoking adjustment, indicating that the adjusted test statistics were less inflated by systemic biases (Supplementary Table 3). Slight increases in genomic inflation factors in AFR REX, AFR AVOID, and AMR REX may be attributable to noise introduced by small sample sizes. Moreover, the standard deviations of the empirical null distributions in EUR and AFR were closer to one after smoking adjustment, also indicating that results were less inflated (Supplementary Table 3). Slight increases in standard deviation in AMR can be attributed to noise introduced by small sample size.

### Replication of previous work

In a previous PGC PTSD EWAS study^11^, there were 11 CpGs that passed the epigenome-wide significance threshold. All 11 passed probe quality control in our sample, and we replicated five of these 11 results (Supplementary Table 4) in our primary analyses. These replications were mostly from *AHRR* and the lowest *p*-values were from the EWAS of AVOID: cg05575921 (*AHRR*, coefficient = −2.06×10^−3^, *p*-value = 1.08×10^−34^), cg21161138 (*AHRR*, coefficient = −6.73×10^−4^, *p*-value = 2.99×10^−21^), cg23576855 (*AHRR*, coefficient = −1.41×10^−3^, *p*-value = 7.58×10^−12^), cg14753356 (intergenic, *IER3/LINC00243*, coefficient = −4.72×10^−4^, *p*-value = 2.87×10^−14^), and cg19719391 (intergenic, *TBC1D19/LOC100506048*, coefficient = 3.94×10^−4^, *p*-value = 2.09×10^−10^). We also identified cg26703534 (*AHRR*) in PCL-Total and AVOID (coefficient = −3.12×10^−4^, *p*-value = 1.82×10^−10^), as well as cg25648203 (*AHRR*) in PCL-Total, REX, AVOID, and HYP (coefficient = −4.66×10^−4^, *p*-value = 1.36×10^−16^), which were not identified in ref. ^11^ but were significant in an earlier study.^13^ One replication survived smoking adjustment: cg05575921 (*AHRR*), coefficient (AVOID) = −7.34×10^−4^, *p*-value = 2.62×10^−8^).

### Correlation analyses

To examine phenotypic and epigenetic differences between lifetime PTSD (PTSD-Dx) and current PTSD symptoms (PCL-Total, REX, AVOID, and HYP), we performed Pearson correlations between the binary phenotype and PCL scores, among the PCL scores, and between their respective methylation coefficients (Supplementary Table 5). All *p*-values were < 2×10^−16^. Phenotypic correlations between PTSD-Dx and PCL scores were 0.59–0.64 in EUR, 0.48–0.54 in AFR, and 0.50-0.56 in AMR. Phenotypic correlations among PCL scores were 0.8–0.9 in all ancestries. Epigenetic correlations between PTSD-Dx and PCL scores were 0.52–0.55 in EUR, 0.29–0.34 in AFR, and 0.29–0.32 in AMR. Epigenetic correlations among PCL scores were 0.8–0.9 in all ancestries.

### Postmortem human brain comparison

To assess whether PTSD-associated methylation signals identified in peripheral blood show cross-tissue convergence with molecular alterations observed in a directly disease-relevant tissue, we compared the significant blood EWAS findings with differentially methylated CpG sites identified in postmortem PTSD brain tissue (subregions of the hippocampus and amygdala; 117 cases, 54 controls; 11.6% female; 3,648 significant CpGs out of 1,136,867 tested) in a previous report.^20^ Five blood and four brain CpGs were located within 1 kb of one another, mapping near *SLC25A34, EXOC2, LIF*, and *NFE2L3* (Fig. 3). At the *LIF* locus, a brain CpG (22:30,639,828, effect size = 4.24, *p*-value = 2.19×10^−5^, FDR-adjusted *p*-value = 0.021) and two blood EWAS loci (cg15722603, 22:30,639,887, coefficient (PCL-Total meta) = −0.0001), lowest *p*-value (PCL-Total meta) = 4.31×10^−8^, and cg23635663, 22:30,639,979, coefficient (AVOID meta) = −0.0001, lowest *p*-value (AVOID meta) = 1.64×10^−9^) were located within 1 kb of one another, and all fell within the same CpG island (22:30,639,600–30,640,100) (Figure 3). Additionally, all were located in a differentially methylated region identified in the blood EWAS in PCL-Total and AVOID (four CpGs, 22:30,639,611−30,639,979, minimum smoothed FDR (PCL-Total) = 1.41×10^−16^, maximum coefficient = −1×10^−4^, mean coefficient = −2.5×10^−5^) (Supplementary Table 6). However, the brain and blood CpGs have opposite directions of effect; the brain CpG is hypermethylated in PTSD, while the blood CpGs are hypomethylated. There was no significant enrichment between the blood and the brain gene sets (OR = 1.1216, enrichment *p*-value = 0.3794). However, 14 of 254 blood EWAS genes (*SLC25A34, HDAC4, RALGDS, NOTCH1, MYO1C, LIF, RIN3, TBCD, PIP5K1C, CREBBP, NFE2L3, LINC00299, ANKRD33B*, and *TULP4*) were present among the 1,208 unique genes annotated to significant PTSD brain CpGs. The overlapping genes are predominantly involved in transcriptional and chromatin regulation, neurodevelopmental and immune signaling, and intracellular trafficking and signaling.^21-23^

**Figure 3.**
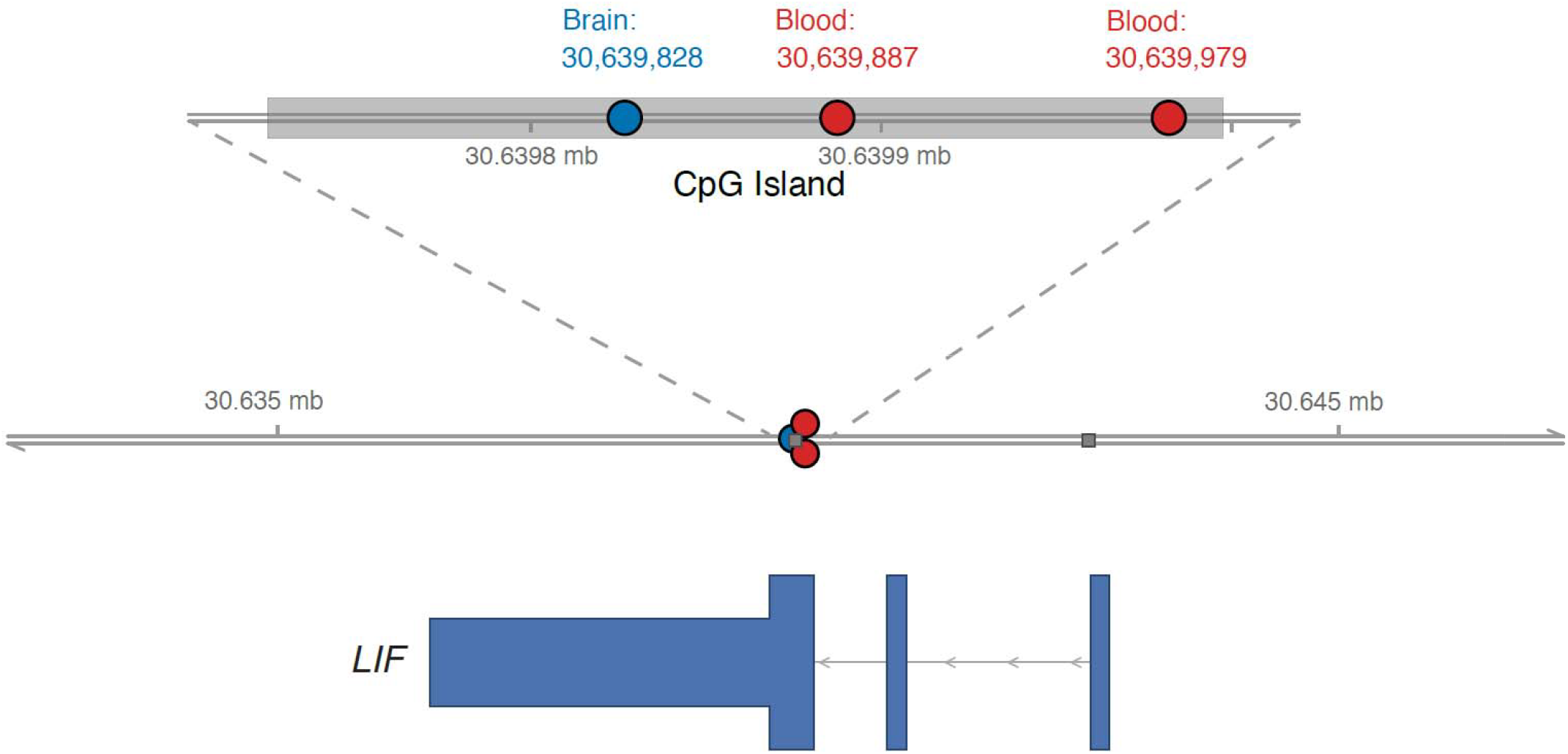
Brain and blood CpG sites localizing to a single CpG island at the *LIF* locus.

### Enrichment analyses

According to Enrichr^24-26^, 10,869 gene sets from 99 libraries, representing publicly available data sources as well as purpose-built datasets^24-26^, were significantly enriched, with FDR < 0.05 for the term associated with the gene set (Supplementary Table 7). From the Allen Brain Atlas, the set of genes downregulated in GABAergic inhibitory interneurons expressing somatostatin (*SST*) and *P4HA3* was enriched (Human Inh L1 *SST P4HA3* down, adjusted *p*-value = 7.85×10^−3^, OR = 12.5, overall score = 135.7).^27^ Five of the six overlapping genes had associated CpG sites with positive effect directions in the EWAS meta-analyses, indicating that a higher PCL-Total score was associated with hypermethylation. (*WDPCP* had a negative direction of effect.) Also from the Allen Brain Atlas, two sets of genes downregulated in astrocytes expressing *FGFR3* were enriched (Hu an Astro L1−6 *FGFR3 AQP1* down, adjusted *p*-value = 9.46×10^−3^, OR = 2.84, overall score = 27.6, and Human Astro L1−6 *FGFR3 PLCG1* down, adjusted *p*-value = 9.46×10^−3^, OR = 2.61, overall score = 25.1), as well as a set of genes downregulated in brain endothelial cells (Human Endo L2−5 *NOSTRIN SRGN* down, adjusted *p*-value = 0.0169, OR = 2.70, overall score = 23.6). Similarly, genes downregulated in interneurons expressing somatostatin and neuropeptide Y in Huntington’s disease (HD) patients vs. control^28^ were enriched (adjusted *p*-value = 2.64×10^−4^, OR = 1.96, overall score = 26.1); this gene set is from HDsigDB, a database of molecular signatures in HD.^28-30^ The GABAergic interneuron gene module, also from HDsigDB, but not derived from HD patients^29^, was also enriched (adjusted *p*-value = 0.0348, OR = 4.85, overall score = 29.9) The set of genes upregulated in astrocytes of HD patients vs. control was enriched (adjusted *p*-value = 3.80×10^−6^, OR = 3.85, overall score = 69.1).

Other significant CpG sites in genes identified in the enrichment analysis include cg21253130, in *FRY*, FRY microtubule-binding protein, which is associated with “neuron projection development.”^31^ This CpG site is also significant in prior EWAS of NO_2_ exposure (a respiratory irritant and potential neurotoxin)^32^, obesity^33^, and type 2 diabetes^33^. Its *p*-value in the EWAS meta-analysis of PCL-Total, with smoking included as a covariate, is 4.52×10^−9^, with a positive direction of effect in all three ancestries.

*CREBBP*, CREB-binding protein, a histone acetyltransferase that acetylates H3K27, has a significant CpG site, cg05194552, in the EWAS meta-analysis of HYP, with *p*-value = 8.62×10^−9^ and a positive effect direction in all three ancestries. Among other roles, *CREBBP* is associated with “[hippocampal] long-term potentiation” (hsa04720).^34^ The cg05194552 probe is significant in EWAS of asthma ^35^ and B-cell acute lymphoblastic leukemia.^36^

*NR3C1*, encoding the glucocorticoid receptor, also has a significant CpG site in the EWAS meta-analysis of PCL-Total, cg22233604 (*p*-value = 6.55×10^−9^, positive direction of effect in all three ancestries). In addition to its endocrine roles, *NR3C1* is associated with “synaptic transmission, glutamatergic” (GO:0035249), “positive regulation of neuron apoptotic process” (GO:0043525), and “astrocyte differentiation” (GO:0048708).^31,37^ The cg22233604 probe is significant in an EWAS of myalgic encephalitis/chronic fatigue syndrome.^38^

*TIMP2*, tissue inhibitor of metalloproteinases 2, has a significant CpG site in primary analyses of PCL-Total, AVOID, and HYP (cg20761853, lowest *p*-value (AVOID) = 5.44×10^−9^, coefficient = 3.65×10^−4^). It is significant in PCL-Total, AVOID, and HYP with smoking included as a covariate (lowest *p*-value (AVOID) = 2.63×10^−9^, coefficient = 3.74×10^−4^), as well as in the meta-analysis of PCL-Total (*p*-value = 3.40×10^−11^, positive direction of effect in all three ancestries). It is highly expressed in the hippocampus and has been found to regulate hippocampal synaptic plasticity and neurogenesis in mice.^39^

### Smoking status stratification

Because lower methylation of CpG sites at *AHRR* has been reliably and very strongly associated with smoking^19^, a previous PTSD EWAS study^13^ examined differences of effect between PTSD and smoking status due to a higher rate of smoking among individuals with PTSD. To test whether effect sizes are statistically significantly different according to smoking status, we stratified by smoking status all the PTSD EWAS analyses that had significant CpGs in *AHRR*, meta-analyzed the results for current, former, and never smokers, and found that they are not significantly different (Supplementary Table 8). None of the 10 CpGs in *AHRR* appearing in our analyses (cg17287155, cg21161138, cg05575921, cg25648203, cg04551776, cg11902777, cg26703534, cg12806681, cg07943658, cg23576855) had significant heterogeneity *p*-values when corrected for multiple testing (*p*-value cutoff = 1×10^−3^, 0.05/50 tests). Within PTSD cases in EUR, there were 30.1% current smokers, 47.1% former smokers, and 22.9% never smokers, for a total of 5,044 individuals (vs. 5,184 when smoking status was not included). Within PTSD controls in EUR, there were 19.6% current smokers, 53.3% former smokers, and 27.1% never smokers, for a total of 18,041 individuals (vs. 19,382 when smoking status was not included). Within PTSD cases in AFR, there were 32.0% current smokers, 40.7% former smokers, and 27.3% never smokers, for a total of 3,024 individuals (vs. 3,095 when smoking status was not included). Within PTSD controls in AFR, there were 29.7% current smokers, 42.5% former smokers, and 27.7% never smokers, for a total of 6,015 individuals (vs. 6,240 when smoking status was not included). Within PTSD cases in AMR, there were 23.6% current smokers, 42.6% former smokers, and 33.8% never smokers, for a total of 1,094 individuals (vs. 1,134 when smoking status was not considered). Within PTSD controls in AMR, there were 18.3% current smokers, 44.7% former smokers, and 37.0% never smokers, for a total of 1,513 individuals (vs. 1,635 when smoking status was not included). The difference in proportion of smoking status between cases and controls was significant in EUR, and for AMR current smokers, but not AFR (*p*-value cutoff = 0.003, 0.05/15 tests) (Supplementary Table 9). Differences among ancestries in proportion of ever smokers (current and former) were significant for EUR vs. AMR and AFR vs. AMR, but not EUR vs. AFR (*p*-value cutoff = 0.004, 0.05/12 tests) (Supplementary Table 9).

### DNA Methylation Adjusted Age

Age at initial blood draw, Horvath DNA methylation age, Levine DNA methylation age, and DunedinPACE rate of aging for PTSD cases and controls in EUR and AFR are provided in Supplementary Table 10. Horvath DNA methylation age was designed to predict chronological age from the methylation state of 353 CpGs and works across tissues.^40^ It was significantly correlated with chronological age in both groups (EUR: N = 24,566, β = 0.893, SE = 0.002, *p*-value < 2×10^−16^, 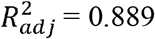; AFR: N = 9,335, β = 0.927, SE = 0.004, *p*-value < 2×10^−16^, 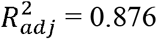). The residuals from this model were retained, representing age adjustment. In the full regression model predicting age adjustment, including covariates of sex, seven cell types, and smoking status, PTSD case status was significantly associated with age adjustment in EUR (*p*-value cutoff = 3.97×10^−4^ = 0.05/126 tests; β = 0.266, SE = 0.0658, *p*-value = 5.46×10^−5^; Supplementary Tables 11, 12). There was no significant association in AFR. Female sex was associated with age adjustment < 0 in EUR (β = −0.750, SE = 0.132, *p*-value = 1.38×10^−8^). Each of the seven cell types was significantly associated with age adjustment in EUR and AFR. In the full regression models (case status and covariates [sex, cell composition, and smoking status]), 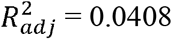 for EUR and 0.0570 for AFR. The mean age adjustment for PTSD cases was not significantly different from zero when sex, cell types, and smoking status were taken as covariates (*p*-value cutoff = 0.00833 = 0.05/6 tests; Supplementary Table 17). DNA methylation age and age adjustment were not analyzed in AMR because of small sample size.

Levine DNA methylation age was designed to predict a composite of clinical biomarkers and mortality risk based on the methylation state of 513 CpGs. It is therefore better at predicting disease and mortality than Horvath.^41^ It was significantly correlated with chronological age in both groups (EUR: N = 19,078, β = 0.888, SE = 0.003, *p*-value < 2×10^−16^, 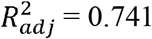; AFR: N = 7,625, β = 0.927, SE = 0.006, *p*-value < 2×10^−16^, 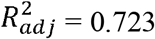). The residuals from this model were retained, representing age adjustment. In the full regression model predicting age adjustment, including covariates of sex, seven cell types, and smoking status, PTSD case status was not significantly associated with age adjustment in EUR or AFR (*p*-value cutoff = 3.97×10^−4^ = 0.05/126 tests; Supplementary Tables 13, 14). When only sex and cell types were taken as covariates, PTSD case status was significantly associated with age adjustment in EUR (β = 0.575, SE = 0.118, p-value = 1.14×10^−6^) but not AFR. Sex was not a significant predictor of age adjustment in EUR or AFR. Each of the seven cell types was significantly associated with age adjustment in EUR and AFR. In the full regression models (case status and covariates [sex, cell composition, and smoking status]), 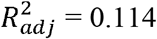for EUR and 0.129 for AFR. The mean age adjustment for PTSD cases was not significantly different from zero in EUR or AFR when sex, cell types, and smoking status were taken as covariates (*p*-value cutoff = 0.00833 = 0.05/6 tests), nor when only sex and cell types were taken as covariates.

DunedinPACE rate of aging was derived from changes in 19 biomarkers over a 20-year period. While Horvath and Levine measure cumulative aging, it measures rate of aging at a point in time.^42^ It was significantly correlated with chronological age in both groups (EUR: N = 24,566, β = 1.62×10^−3^, SE = 5.92×10^−5^, *p*-value < 2×10^−16^, 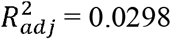; AFR: N = 9, 335, β = 3.04×10^−3^, SE = 1.07×10^−4^, *p*-value < 2×10^−16^, 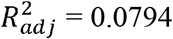). The residuals from this model were retained, representing change in rate of aging (age acceleration/deceleration). In the full regression model predicting change in rate of aging, including covariates of sex, seven cell types, and smoking status, PTSD case status was significantly associated with accelerated aging in EUR (*p*-value cutoff = 3.97×10^−4^ = 0.05/126 tests; β = 0.00843, SE = 0.00174, *p*-value = 1.29×10^−6^; Supplementary Tables 15, 16), but not in AFR. Sex was not associated with change in rate of aging in EUR, but female sex was significantly associated with accelerated aging in AFR (β = 0.0175, SE = 0.00402, *p*-value = 1.29×10^−5^). Each of the seven cell types was significantly associated with accelerated aging in EUR and AFR. In the full regression models (case status and covariates [sex, cell composition, and smoking status]), 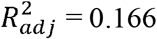 for EUR and 0.229 for AFR. The mean epigenetic age acceleration for PTSD cases was not significantly greater than zero when sex, cell types, and smoking status were taken as covariates (*p*-value cutoff = 0.00833 = 0.05/6 tests; Supplementary Table 17). When only sex and cell types were taken as covariates, the mean epigenetic age acceleration was significantly different from zero (mean = 7.04×10^−3^, *p*-value = 5.74×10^−6^) (Supplementary Table 17).

## Methods

### Cohorts and PTSD definitions

**Table 1:** Demographics of PTSD traits.

|  | Sample Size | Score (1<sup>st</sup> quartile, 2<sup>nd</sup> quartile, 3<sup>rd</sup> quartile) or numbers of cases and mean + SD | Average Age | Males/Females (percentage) |
| --- | --- | --- | --- | --- |
|  | EUR |  |  |  |
| PCL-Total | 17,674 | (19, 24.8, 37) | 69.3 | 95.3% males, 4.7% females |
| Re-experiencing | 17,674 | (5, 6, 10) | 69.3 | 95.3% males, 4.7% females |
| Avoidance | 17,674 | (7, 10, 15.2) | 69.3 | 95.3% males, 4.7% females |
| Hyperarousal | 17,674 | (6, 8, 12) | 69.3 | 95.3% males, 4.7% females |
| Lifetime PTSD | 24,566 | 21.1% cases and 78.9% controls | 68.1 | 95.4% males, 4.6% female |
|  | AFR |  |  |  |
| PCL-Total | 3,430 | (21, 30, 48) | 63.3 | 89.6% males, 10.4% females |
| Re-experiencing | 3,430 | (5, 8, 14) | 63.3 | 89.6% males, 10.4% females |
| Avoidance | 3,430 | (8, 12, 20) | 63.3 | 89.6% males, 10.4% females |
| Hyperarousal | 3,430 | (6, 9, 15) | 63.3 | 89.6% males, 10.4% females |
| Lifetime PTSD | 9,335 | 33.2% cases and 66.8% controls | 60.8 | 89.8% males, 10.2% females |
|  | AMR |  |  |  |
| PCL-Total | 1,037 | (21, 33, 52) | 63.1 | 90.6% males, 9.4% females |
| Re-experiencing | 1,037 | (6, 9, 15) | 63.1 | 90.6% males, 9.4% females |
| Avoidance | 1,037 | (8, 13, 22) | 63.1 | 90.6% males, 9.4% females |
| Hyperarousal | 1,037 | (7, 10, 16) | 63.1 | 90.6% males, 9.4% females |
| Lifetime PTSD | 2,769 | 41.0% cases and 59.0% controls | 58.1 | 91.6% males, 8.4% females |

We considered five PTSD traits—two diagnostic traits (PCL-Total and Lifetime PTSD) and three symptom subdomains (REX, AVOID, and HYP). PCL-Total is the PCL-C for DSM-IV used to assess PTSD symptoms over the past month by self-report; scores range from 17 to 85.^16^ The other three scores are subscales of the PCL-C. REX is the sum of questions 1–5; the range is from 5 to 25. AVOID is the sum of questions 6–12; the range is from 7 to 35. HYP is the sum of questions 13–17; the range is from 5 to 25. The Lifetime PTSD variable used the case and control outputs from lasso regression with cross-validation based on manual chart review as the “gold standard.”^17^ The lasso algorithm used EHR data to classify Lifetime PTSD by selecting the category with the highest probability from the following: case = PTSD case, possible = possible PTSD case, and control = PTSD control. We used only the cases and controls, excluding the “possible” category from analyses. Ancestry was aggregated into population groups—EUR, AFR, and AMR—based on genetic similarity to the 1000 Genomes Project reference panels.^43^ For PCL-Total, REX, AVOID, and HYP, there were a total of 22,141 samples, 17,674 for EUR, 3,430 for AFR, and 1,037 for AMR. For Lifetime PTSD, there were 9,413 cases and 27,257 controls, with 5,184 cases and 19,382 controls for EUR, 3,095 cases and 6,240 controls for AFR, and 1,134 cases and 1,635 controls for AMR. Demographics for the group of subjects with smoking status available are in Supplementary Table 18.

### DNA methylation quality control

The following quality control was done by the MVP epigenetics group.^44^ Samples for the MVP methylation data were selected based on enrichment of phenotype and advanced age because there is a higher prevalence of diseases relevant to veterans in older people. Raw fluorescence intensity data (IDAT files) were converted to methylation beta values. SeSAMe was used for processing the methylation data and for quality control, along with *P*-value with Out Of Band probes for Array Hybridization (pOOBAH).^45^ In the probe quality control, a given probe was removed if it had a pOOBAH detection *p*-value ≥ 0.05 in > 10% of samples. There were 865,918 total probes on the Illumina Infinium Methylation EPIC v1 array, which was run on 45,500 whole-blood samples from the MVP cohort to generate DNA methylation profiles. After removing non-CpG probes, 862,927 probes were retained. Probes with pOOBAH failure rate in > 10% of samples were removed, leaving 836,196 probes. Technically unreliable probes were removed due to poor mapping quality, poor titration correlation, or color channel switching, leaving 768,569 probes. The final number of CpG probes in the MVP data release was 768,569^44^, yielding a Bonferroni-corrected *p*-value threshold of 6.51×10^−8^ (0.05/768,569) for epigenome-wide significance.

### Epigenome-wide association analysis

The meffil R package^46^ was used to fit the EWAS analyses, with methylation betas as outcome, using as covariates age, sex, Houseman cell type proportion^47^, 20 genotype principal components, 20 technical principal components, scanner ID, and duration of sample storage. Cell types included B cells, CD4+ T cells, CD8+ T cells, eosinophils, monocytes, neutrophils, and natural killer cells. In the sensitivity analyses performed for all traits, smoking status was added as a covariate. Smoking status was defined using the lasso algorithm, which returns the probability of being a never, former, or current smoker; the category with the highest probability was selected as the smoking value for each individual.^48^ The R package BACON^18^ was used to control inflation. It constructs an empirical null distribution using a Gibbs sampling algorithm, fitting a three-component normal mixture on *z*-scores. The genomic inflation factors were closer to one after doing so (Supplementary Table 3). Epigenome-wide significance was defined as a BACON-and Bonferroni-corrected *p*-value < 6.51×10^−8^ (0.05/768,569 tests). To generate a list of significant CpGs and their mapped genes for blood-brain comparison and enrichment analyses, we used METAL^49^ to perform sample-size-weighted multi-ancestry meta-analyses of our EWAS results with smoking included as a covariate for all five PTSD traits; effective sample size was used for Lifetime PTSD. Additionally, we used cor.test() in base R to perform a Pearson correlation among phenotypes, and among methylation coefficients, for PTSD-Dx and PCL scores, to assess for differences in methylation between lifetime PTSD diagnosis and current PTSD symptoms (Supplementary Table 5). We also performed a differentially methylated region (DMR) analysis using the R package DMRcate (Supplementary Tables 6, 19, 20).^50^

### Smoking status stratification

To assess whether effect sizes for smoking-associated CpGs in *AHRR* vary according to smoking status, we conducted separate EWASes of current, former, and never smokers in all five PTSD traits, and then meta-analyzed current, former, and never smokers for each trait, using METAL to generate heterogeneity statistics (Supplementary Table 8).

### DNA Methylation Adjusted Age

Horvath DNA methylation age estimates were calculated using 353 CpG sites in 8,000 samples, across 51 healthy tissues and cell types.^40^ DunedinPACE, a DNA methylation estimate of the rate of aging, was calculated using 173 CpG sites across 19 organ-system biomarkers in 1,037 participants.^42^ Levine PhenoAge estimates were calculated using 513 CpG sites in 9,926 NHANES III participants.^41,51^ We regressed each measure of DNA methylation age or rate of aging onto chronological age at time of initial blood draw and retained the residuals to reflect age adjustment (for Horvath and Levine) and change in rate of aging (for DunedinPACE). To investigate the relationship between age adjustment or change in rate of aging and PTSD case status (EUR: 5,184 cases and 19,382 controls; AFR: 3,095 cases and 6,240 controls), we regressed the residuals onto case status, with covariates of genetically defined sex, seven cell types, and smoking status (Supplementary Tables 11, 12, 13, 14, 15, 16). For Lifetime PTSD, we regressed Horvath age onto chronological age, with covariates of sex, cell type proportion, and smoking status, did the same for Levine PhenoAge and DunedinPACE rate of aging, and retained the residuals. Then we performed a one-sample *t*-test of the residuals for PTSD cases only to test whether the mean age adjustment or change in rate of aging for PTSD cases was significantly different from zero (Supplementary Table 17).

### Brain CpG and blood EWAS overlap

Differentially methylated CpG sites associated with PTSD were identified using targeted methyl-seq data from human postmortem brain, spanning subregions of the amygdala (medial, basolateral, central) and hippocampus (subiculum, dentate gyrus, CA subfields), as reported previously.^20^ Significant CpGs were called with the DSS package from Bioconductor^52^, fitting a beta-binomial model with an arc-sine link function, with diagnosis group as the variable of interest and brain subregion, age, sex, postmortem interval, tissue source, smoking status, neuronal proportion, and ancestry included as covariates. By adjusting for brain subregion, we conducted a cross-region analysis designed to identify PTSD-associated methylation differences while accounting for regional variation in DNA methylation. This approach increased statistical power by leveraging data across all six brain regions and prioritized signals with evidence of association across PTSD-relevant subregions of the amygdala and hippocampus. CpGs with a Benjamini-Hochberg-adjusted FDR < 0.05 were considered significant and annotated to the nearest gene. Based on the current blood-based analyses and the previously reported brain-based analyses^20^, overlap with blood EWAS results was evaluated at both the CpG and gene levels. At the CpG level, brain CpG coordinates (hg38) were converted to build 37 using liftOver^53^, then compared with blood EWAS CpG coordinates to identify exact matches and loci within ±1 kb, using the findOverlaps() function from the GenomicRanges R package.^54^ At the gene level, the set of genes annotated to significant brain CpGs was tested for enrichment for genes from the blood EWAS results using the R package GeneOverlap^55^ and Fisher’s exact test.

### Enrichment Analyses

We performed gene set enrichment analysis with Enrichr^24-26^ (Supplementary Table 7). Enrichr uses Fisher’s exact test, calculates the odds ratio (OR) to determine whether genes from the input list appear in a gene set more often than would be expected by chance, and applies the Benjamini-Hochberg correction to account for multiple testing across all tests performed in the run. It then calculates an overall enrichment score equal to the negative log of the *p*-value multiplied by the OR.^24-26^ The input gene list of 227 genes included one gene from a trans-ancestry meta-analysis of Lifetime PTSD, 151 additional genes from a trans-ancestry meta-analysis of PCL-Total, 21 additional genes from a trans-ancestry meta-analysis of REX, 53 additional genes from a transancestry meta-analysis of AVOID, and two additional genes from a trans-ancestry meta-analysis of HYP. All meta-analyzed EWAS included smoking status as a covariate. Significant CpG sites from the meta-analyses were mapped to genes using the R package IlluminaHumanMethylationEPICanno.ilm10b4.hg19.^56^ Significant CpG sites of interest, those mapping to genes found both in our input gene list and in brain- or PTSD-relevant gene sets, were annotated using CpG Atlas.^37^

## Discussion

In this study, we performed EWAS of PTSD across multiple ancestries using validated algorithmic and EHR diagnoses, quantitative PCL scores, and diagnosis subphenotypes. We include 9,413 cases with a total of 36,670 participants for Lifetime PTSD diagnoses; in the previous largest PTSD EWAS, there were 2,156 PTSD cases with a total of 5,077 participants. This U.S. military sample, though ancestrally diverse, is also comparatively homogeneous with regard to sex (90% male), healthcare access, and shared military exposures. We include 22,141 individuals who completed the PCL-C to give scores for PCL-Total, REX, AVOID, and HYP, to maximize power for EWAS based on quantitative total severity scores and symptom domain subscales. In the PTSD EWAS of blood DNA methylation levels, we identified a total of 272 unique CpG sites associated with PTSD and 217 genes, replicating five CpG sites that were identified in a prior meta-analysis of PTSD EWAS. Three of the five replicated CpG sites all map to *AHRR*, and thus may reflect smoking status, and two are intergenic. The identified PTSD-associated CpGs had epigenome-wide significance between 1.08×10^−34^ ≤ *p* < 6.51×10^−8^. The most significant CpGs had their lowest *p*-values in AVOID and were mapped to *AHRR* (cg05575921, coefficient = −2.06×10^−3^, *p*-value = 1.08×10^−34^; cg21161138, coefficient = −6.73×10^−4^, *p*-value = 2.99×10^−21^), *F2RL3* (cg03636183, coefficient = −8.27×10^−4^, *p*-value = 5.06×10^−22^), or *RARA* (cg17739917, coefficient = −7.73×10^−4^, *p*-value = 7.16×10^−20^), or were intergenic (cg21566642, coefficient = −1.09×10^−3^, *p*-value = 2.40×10^−21^; cg01940273, coefficient = −6.86×10^−4^, *p*-value = 1.02×10^−19^). *AHRR* is well-known to be affected by smoking.^19,57^ These genes have been implicated in smoking (*AHRR, F2RL3*)^19,57,58^, inflammation and immune disorders (*GPR15, RARA, ITGAL*)^59-61^, regulation of hematopoietic cell differentiation (*ASB2*)^62^, myocardial infarction and platelet function (*F2RL3*)^63^, cancer/tumor progression and drug resistance (*PRSS23, RARA, SLC1A5, ITGAL*)^64-66^, and metabolic and bone disorders (*SLC1A5*)^67^, and have also been identified as potential targets for gastric or breast cancer therapy (*DENND2D*).^68^

A challenge in the interpretation of EWAS results is relating findings in peripheral blood to methylation patterns in the brain, the presumed locus of the most relevant biology. There is often correspondence in epigenetic findings between brain and peripheral blood.^69^ Genes associated with significant CpGs in both the blood and the previously reported brain analyses^20^ were *SLC25A34, HDAC4, RALGDS, NOTCH1, MYO1C, LIF, RIN3, TBCD, PIP5K1C, CREBBP, NFE2L3, LINC00299, ANKRD33B*, and *TULP4*. The protein products of these genes affect aspects of metabolic regulation—including mitochondrial substrate transport (*SLC25A34*)^70^, vesicle exocytosis (*EXOC2*)^71^, cytokine signaling *(LIF*)^72^, and stress-responsive transcription (*NFE2L3*).^73^ In a CpG island intronic to *LIF*, leukemia inhibitory factor interleukin 6 family cytokine, we found a brain CpG and a blood CpG (cg23635663) 151 bp apart. *LIF* is neuroprotective, anti-inflammatory, and promotes neural stem cell maintenance and tissue repair.^74^ At *EXOC2*, the blood and brain CpGs were also separated by less than 1 kb, providing a second example of local cross-tissue convergence. Although these genes do not constitute a recognized molecular pathway, their identification in EWAS of two biologically distinct tissues provides evidence of cross-tissue convergence and supports their prioritization for further investigation in PTSD.

Enrichment analyses revealed enrichments in multiple brain cell types, including somatostatin interneurons, which accords with recent scRNAseq analyses of brain tissue of individuals with PTSD^75^, GABAergic interneurons generally, astrocytes, and brain endothelial cells. The involvement of the glucocorticoid receptor points to the role of the stress response in PTSD.^76-78^ The CpG site we identify in *NR3C1*, cg22233604 (5:1472729377, meta-analyses of PCL-Total and AVOID, lower *p*-value = 4.68×10^−10^ (AVOID), positive coefficient in all three ancestries), lies in the body of the gene, not in the promoter region; the hypermethylation of this site in our results is therefore hard to interpret in terms of likely functional outcome.

Our meta-analyses of EWASes stratified by smoking status showed no significant differences in effect size for the 10 CpGs that we tested in *AHRR*. This contrasts with findings reported previously^13^ that associations between *AHRR* methylation and PTSD were most prominent among non-smokers and absent among smokers, leading to their interpretation that *AHRR* methylation reflected PTSD-related biology in addition to smoking exposure; in that report, the *AHRR* association was interpreted to reflect PTSD in addition to smoking. In contrast, our results suggest that observed associations between PTSD and *AHRR* methylation reflect smoking-related effects rather than PTSD-specific epigenetic changes. Cohort characteristics differed between studies: former smokers comprised the majority of both PTSD cases and controls in the MVP, whereas the prior study sample had a higher prevalence of current smoking among PTSD cases. Our sample sizes were also much larger, allowing us to stratify three smoking statuses of “current,” “former,” and “never,” whereas they had two categories of smokers and non-smokers. Our conclusions regarding *AHRR* and PTSD thus differ substantially from previous reports. We conclude that the observed association most likely reflects smoking, not PTSD.

We examined algorithmically defined PTSD case status as a predictor of residuals derived from regressing methylation age or rate of aging onto chronological age. PTSD case status was significantly associated with excess aging or age acceleration in EUR ancestry when controlling for sex, chronological age, cell type proportion, and smoking status (Horvath β = 0.266, *p*-value = 5.46×10^−5^; DunedinPACE β = 8.43×10^−3^, *p*-value = 1.29×10^−6^) (Supplementary Tables 11, 12, 13, 14, 15, 16). The association was not statistically significant in AFR and not tested in AMR. These results are consistent with prior reports demonstrating significant age acceleration associated with PTSD.^79,80^ A meta-analysis of mostly military veteran cohorts (n = 1,367) found that Horvath age residuals were significantly associated with an interaction term between PTSD onset or change in symptom severity and Horvath age residuals at an earlier time point (new onset β = 0.16, adjusted *p*-value = 0.03; change in severity β = 0.24, *p*-value = 0.05).^79^ A meta-analysis primarily of veterans (n = 2,186), found significant associations between excess aging per the Hannum clock^81^, but not the Horvath.^80^

Our study has limitations. Our U.S. military veteran sample was mostly male (95.3% in EUR, 89.6% in AFR, 90.6% in AMR), and findings in females may differ. The other side of this limitation is that the higher homogeneity by sex may have improved statistical power, albeit for findings in males. While our study has comparatively good representation of non-EUR individuals (67.0% EUR, 25.5% AFR, and 7.6% AMR for Lifetime PTSD; 79.8% EUR, 15.5% AFR, and 4.7% AMR for PCL scores), the lower number of non-EUR limited discovery in those populations. Study of blood rather than brain is a limitation for linking epigenetic variants to regulation of genes in the brain, but an advantage for possible future clinical applications. For any clinical use of these data in living patients, e.g., for treatment personalization, results are useful only if they can be obtained in readily-accessible tissues. Further, this study was cross-sectional, with a mean age of blood draw of approximately 65.4 years. Most participants were diagnosed with PTSD many years before the measurement of methylation levels by array, i.e., we likely detected chronic rather than acute methylation differences.

Correlation analysis of methylation betas revealed correlation coefficients of 0.8–0.9 between REX, AVOID, and HYP (measures of current PTSD symptoms) in all ancestries, but only approximately 0.5 (in EUR) between PTSD-Dx and PCL scores, suggesting that there are epigenetic differences between current and lifetime exposures; this deserves further study.

PTSD is essentially linked to environmental exposure, so epigenetic mechanisms may be particularly relevant because they can yield persistent molecular correlates of trauma and stress. Epigenetic variation may help explain why individuals with similar trauma exposure differ in PTSD susceptibility and symptom severity beyond differences attributable to genetic risk alone. Our work represents the largest-to-date epigenetic analysis of PTSD, with the identification of many novel epigenetically-modified loci associated with this trauma-related trait and the discovery of new biology. Our sample was less heterogenous, across multiple dimensions, than prior meta-analyzed samples. In the largest single-cell genomics analysis of PTSD in the human brain^75^, genes and pathways associated with PTSD included immune and neuroinflammatory mechanisms, which accords with our EWAS study’s interpretations as well as those of the prior literature.^11-13^ We identify convergence between blood and postmortem brain methylation findings and implicate brain-relevant cellular programs together with accelerated DNA methylation aging. These findings substantially extend the molecular characterization of PTSD while illustrating the importance of distinguishing PTSD-associated epigenetic variation from correlated environmental exposures, e.g., smoking. This work may eventually have clinical utility, by allowing the identification of novel pharmacological targets for PTSD, a disorder for which there presently are few pharmacological treatments—and those that exist are not highly effective.

## Data availability

All MVP and meta-analysis summary statistics are made available through dbGAP, https://dbgap.ncbi.nlm.nih.gov, under accession number phs001672.

## Code availability

Code for all software packages used in this analysis is publicly available via the citations for each method.

## Acknowledgements

This research is based in part on data from the Million Veteran Program, Office of Research and Development, Veterans Health Administration, and was supported by MVP000 as well as award I01BX006482-05. More details regarding this consortium are available in the Supplementary Information. Further support was provided by NIH (grants R01MH133728 to J.G. and M.B.S., and 5R01DA058862 to J.G.). D.F.L. is supported by a Career Development Award CDA-2 from the Veterans Affairs Office of Research and Development (1IK2BX005058-01A2). This publication does not represent the views of the Department of Veterans Affairs or the United States Government.

## Author Contributions

J.G., C.D., and D.F.L. designed the study. S.B., J.G., C.D., and M.J.G. drafted the manuscript. D.F.L. provided ongoing feedback and refinement of the analytical plan. C.D., J.G., M.B.S., K.M.H., and D.F.L. discussed and refined the phenotype in MVP. D.F.L., S.B., C.D., and J.G. discussed and refined analytic plans. S.B., C.D., C.O., and J.C. conducted original analyses. D.F.L., J.G., and M.J.G. supervised original analyses. J.G. supervised the work and, with M.B.S., obtained funding and provided administrative support. All authors critically evaluated and revised the manuscript.

## References

1. American Psychiatric Association. Diagnostic and Statistical Manual of Mental Disorders: DSM-IV, (American Psychiatric Association, Washington, DC, 1994).

2. Zen, A.L., Whooley, M.A., Zhao, S. & Cohen, B.E. Post-traumatic stress disorder is associated with poor health behaviors: findings from the heart and soul study. Health Psychol 31, 194–201 (2012).

3. Jacobsen, L.K., Southwick, S.M. & Kosten, T.R. Substance use disorders in patients with posttraumatic stress disorder: a review of the literature. Am J Psychiatry 158, 1184–90 (2001).

4. Kessler, R.C. et al. Trauma and PTSD in the WHO World Mental Health Surveys. Eur J Psychotraumatol 8, 1353383 (2017).

5. Koenen, K.C. et al. Posttraumatic stress disorder in the World Mental Health Surveys. Psychol Med 47, 2260–2274 (2017).

6. Gelernter, J. et al. Genome-wide association study of post-traumatic stress disorder reexperiencing symptoms in >165,000 US veterans. Nat Neurosci 22, 1394–1401 (2019).

7. Stein, M.B. et al. Genome-wide association analyses of post-traumatic stress disorder and its symptom subdomains in the Million Veteran Program. Nat Genet 53, 174–184 (2021).

8. Nievergelt, C.M. et al. Genome-wide association analyses identify 95 risk loci and provide insights into the neurobiology of post-traumatic stress disorder. Nat Genet 56, 792–808 (2024).

9. Bestor, T.H. The DNA methyltransferases of mammals. Hum Mol Genet 9, 2395–402 (2000).

10. Zannas, A.S., Provencal, N. & Binder, E.B. Epigenetics of Posttraumatic Stress Disorder: Current Evidence, Challenges, and Future Directions. Biol Psychiatry 78, 327–35 (2015).

11. Katrinli, S. et al. Epigenome-wide association studies identify novel DNA methylation sites associated with PTSD: a meta-analysis of 23 military and civilian cohorts. Genome Med 16, 147 (2024).

12. Logue, M.W. et al. An epigenome-wide association study of posttraumatic stress disorder in US veterans implicates several new DNA methylation loci. Clin Epigenetics 12, 46 (2020).

13. Smith, A.K. et al. Epigenome-wide meta-analysis of PTSD across 10 military and civilian cohorts identifies methylation changes in AHRR. Nat Commun 11, 5965 (2020).

14. Philibert, R.A., Beach, S.R. & Brody, G.H. Demethylation of the aryl hydrocarbon receptor repressor as a biomarker for nascent smokers. Epigenetics 7, 1331–8 (2012).

15. Jones, G.T. et al. A DNA Methylation Marker, cg05575921 (AHRR), Outperforms Self-Reported Smoking Exposure for Its Association With Cardiovascular Disease Prevalence. Nicotine Tob Res (2025).

16. Weathers, F.W., Litz, B.T., Herman, D.S., Huska, J.A., and Keane, T.M. The PTSD Checklist (PCL): Reliability, Validity, and Diagnostic Utility. in Annual Meeting of the International Society for Traumatic Stress Studies (San Antonio, TX, 1993).

17. Harrington, K.M. et al. Validation of an Electronic Medical Record-Based Algorithm for Identifying Posttraumatic Stress Disorder in U.S. Veterans. J Trauma Stress 32, 226–237 (2019).

18. van Iterson, M., van Zwet, E.W., Consortium, B. & Heijmans, B.T. Controlling bias and inflation in epigenome- and transcriptome-wide association studies using the empirical null distribution. Genome Biol 18, 19 (2017).

19. Pospiech, E. et al. DNA methylation at AHRR as a master predictor of smoke exposure and a biomarker for sleep and exercise. Clin Epigenetics 16, 147 (2024).

20. Li, H. et al. Mapping DNA Methylation Signatures to Identify Epigenetic Variation Across Subcortical Regions of the Human Posttraumatic Stress Disorder Brain. Biol Psychiatry 100, 76–87 (2026).

21. Sarshad, A.A. et al. Glycogen synthase kinase (GSK) 3beta phosphorylates and protects nuclear myosin 1c from proteasome-mediated degradation to activate rDNA transcription in early G1 cells. PLoS Genet 10, e1004390 (2014).

22. Bauer, S. & Patterson, P.H. Leukemia inhibitory factor promotes neural stem cell self-renewal in the adult brain. J Neurosci 26, 12089–99 (2006).

23. Flex, E. et al. Biallelic Mutations in TBCD, Encoding the Tubulin Folding Cofactor D, Perturb Microtubule Dynamics and Cause Early-Onset Encephalopathy. Am J Hum Genet 99, 962–973 (2016).

24. Chen, E.Y. et al. Enrichr: interactive and collaborative HTML5 gene list enrichment analysis tool. BMC Bioinformatics 14, 128 (2013).

25. Kuleshov, M.V. et al. Enrichr: a comprehensive gene set enrichment analysis web server 2016 update. Nucleic Acids Res 44, W90–7 (2016).

26. Xie, Z. et al. Gene Set Knowledge Discovery with Enrichr. Curr Protoc 1, e90 (2021).

27. Bakken, T.E. et al. Comparative cellular analysis of motor cortex in human, marmoset and mouse. Nature 598, 111–119 (2021).

28. Lee, H. et al. Cell Type-Specific Transcriptomics Reveals that Mutant Huntingtin Leads to Mitochondrial RNA Release and Neuronal Innate Immune Activation. Neuron 107, 891–908 e8 (2020).

29. Winden, K.D. et al. The organization of the transcriptional network in specific neuronal classes. Mol Syst Biol 5, 291 (2009).

30. Aaronson, J. et al. HDinHD: A Rich Data Portal for Huntington’s Disease Research. J Huntingtons Dis 10, 405–412 (2021).

31. Ashburner, M. et al. Gene ontology: tool for the unification of biology. The Gene Ontology Consortium. Nat Genet 25, 25–9 (2000).

32. de, F.C.L.A.J. et al. Long-term Air Pollution Exposure, Genome-wide DNA Methylation and Lung Function in the LifeLines Cohort Study. Environ Health Perspect 126, 027004 (2018).

33. Kirchner, H. et al. Altered DNA methylation of glycolytic and lipogenic genes in liver from obese and type 2 diabetic patients. Mol Metab 5, 171–183 (2016).

34. Kanehisa, M., Furumichi, M., Sato, Y., Matsuura, Y. & Ishiguro-Watanabe, M. KEGG: biological systems database as a model of the real world. Nucleic Acids Res 53, D672–D677 (2025).

35. Nicodemus-Johnson, J. et al. DNA methylation in lung cells is associated with asthma endotypes and genetic risk. JCI Insight 1, e90151 (2016).

36. Gabriel, A.S. et al. Epigenetic landscape correlates with genetic subtype but does not predict outcome in childhood acute lymphoblastic leukemia. Epigenetics 10, 717–26 (2015).

37. Armstrong, J.F. et al. CpG Atlas: A centralized multi-layer database and AI interface for DNA methylation research. bioRxiv (2026).

38. Trivedi, M.S. et al. Identification of Myalgic Encephalomyelitis/Chronic Fatigue Syndrome-associated DNA methylation patterns. PLoS One 13, e0201066 (2018).

39. Ferreira, A.C. et al. Neuronal TIMP2 regulates hippocampus-dependent plasticity and extracellular matrix complexity. Mol Psychiatry 28, 3943–3954 (2023).

40. Horvath, S. DNA methylation age of human tissues and cell types. Genome Biol 14, R115 (2013).

41. Levine, M.E. et al. An epigenetic biomarker of aging for lifespan and healthspan. Aging (Albany NY) 10, 573–591 (2018).

42. Belsky, D.W. et al. DunedinPACE, a DNA methylation biomarker of the pace of aging. Elife 11 (2022).

43. 1000 Genomes Project Consortium et al. A global reference for human genetic variation. Nature 526, 68–74 (2015).

44. Schreiner, P.A., Markianos, K., Francis, M., Despard, B., Gorman, B. R., Said, I., Dong, F., Gautam, S., Dochtermann, D., Shi, Y., Devineni, P., Kirkpatrick, C., Khazanov, N., Moser, J., VA Million Veteran Program, Huang, G. D., Muralidhar, S., Tsao, P. S., Pyarajan, S. Methylation profiling in the Million Veteran Program: design, quality control, and smoking-associated epigenetic signatures. medRxiv (2026).

45. Zhou, W., Triche, T.J., Jr., Laird, P.W. & Shen, H. SeSAMe: reducing artifactual detection of DNA methylation by Infinium BeadChips in genomic deletions. Nucleic Acids Res 46, e123 (2018).

46. Min, J.L., Hemani, G., Davey Smith, G., Relton, C. & Suderman, M. Meffil: efficient normalization and analysis of very large DNA methylation datasets. Bioinformatics 34, 3983–3989 (2018).

47. Houseman, E.A. et al. DNA methylation arrays as surrogate measures of cell mixture distribution. BMC Bioinformatics 13, 86 (2012).

48. Song, R.J. et al. Development of an Electronic Health Record-Based Algorithm for Smoking Status Using the Million Veteran Program (MVP) Cohort Survey Response. Circulation 134(2016).

49. Willer, C.J., Li, Y. & Abecasis, G.R. METAL: fast and efficient meta-analysis of genomewide association scans. Bioinformatics 26, 2190–1 (2010).

50. Peters, T.J. et al. De novo identification of differentially methylated regions in the human genome. Epigenetics Chromatin 8, 6 (2015).

51. Burt, V.L. & Harris, T. The third National Health and Nutrition Examination Survey: contributing data on aging and health. Gerontologist 34, 486–90 (1994).

52. Park, Y. & Wu, H. Differential methylation analysis for BS-seq data under general experimental design. Bioinformatics 32, 1446–53 (2016).

53. Genovese, G. et al. BCFtools/liftover: an accurate and comprehensive tool to convert genetic variants across genome assemblies. Bioinformatics 40 (2024).

54. Lawrence, M. et al. Software for computing and annotating genomic ranges. PLoS Comput Biol 9, e1003118 (2013).

55. Shen, L. GeneOverlap: Test and visualize gene overlaps. R package version 1.26.0 edn (2020).

56. Hansen, K.D. IlluminaHumanMethylationEPICanno.ilm10b4.hg19: Annotation for Illumina’s EPIC methylation arrays. 0.6.0 edn (2017).

57. Chybowska, A.D. et al. A blood- and brain-based EWAS of smoking. Nat Commun 16, 3210 (2025).

58. Zhang, Y., Yang, R., Burwinkel, B., Breitling, L.P. & Brenner, H. F2RL3 methylation as a biomarker of current and lifetime smoking exposures. Environ Health Perspect 122, 131–7 (2014).

59. Cui, J. et al. GPR15-guided CD8(+) T regulatory cells control intestinal inflammation. Nature 656, 1003–1012 (2026).

60. Larange, A. et al. A regulatory circuit controlled by extranuclear and nuclear retinoic acid receptor alpha determines T cell activation and function. Immunity 56, 2054–2069 e10 (2023).

61. Lin, F. et al. Magnesium-related gene ITGAL: a key immunotherapy predictor and prognostic biomarker in pan-cancer. Front Pharmacol 15, 1464830 (2024).

62. Wang, J., Muntean, A.G. & Hess, J.L. ECSASB2 mediates MLL degradation during hematopoietic differentiation. Blood 119, 1151–61 (2012).

63. Corbin, L.J. et al. Epigenetic Regulation of F2RL3 Associates With Myocardial Infarction and Platelet Function. Circ Res 130, 384–400 (2022).

64. Xiong, J.X. et al. Targeting PRSS23 with tipranavir induces gastric cancer stem cell apoptosis and inhibits growth of gastric cancer via the MKK3/p38 MAPK-IL24 pathway. Acta Pharmacol Sin 45, 405–421 (2024).

65. Zhang, G. et al. Elevated SLC1A5 associated with poor prognosis and therapeutic resistance to transarterial chemoembolization in hepatocellular carcinoma. J Transl Med 22, 543 (2024).

66. Zhang, R. et al. ITGAL expression in non-small-cell lung cancer tissue and its association with immune infiltrates. Front Immunol 15, 1382231 (2024).

67. Sharma, D., Yu, Y., Shen, L., Zhang, G.F. & Karner, C.M. SLC1A5 provides glutamine and asparagine necessary for bone development in mice. Elife 10 (2021).

68. Chang, C.F. et al. Unveiling DENND2D as a Novel Prognostic Biomarker for Prostate Cancer Recurrence: From Gene to Prognosis. Biomedicines 13 (2024).

69. Braun, P.R. et al. Genome-wide DNA methylation comparison between live human brain and peripheral tissues within individuals. Transl Psychiatry 9, 47 (2019).

70. Karavaeva, I. et al. Mitochondrial carrier SLC25A34 links clock, diet, and temperature control of adipocyte lipid cycling. Science 394, eadz4797 (2026).

71. Van Bergen, N.J. et al. Mutations in the exocyst component EXOC2 cause severe defects in human brain development. J Exp Med 217 (2020).

72. Nicola, N.A. & Babon, J.J. Leukemia inhibitory factor (LIF). Cytokine Growth Factor Rev 26, 533–44 (2015).

73. Yaker, L. et al. NFE2L3 regulates inflammation and oxidative stress-related genes in the colon. Biochim Biophys Acta Mol Cell Res 1872, 119987 (2025).

74. Wang, J. et al. Leukemia inhibitory factor, a double-edged sword with therapeutic implications in human diseases. Mol Ther 31, 331–343 (2023).

75. Hwang, A. et al. Single-cell transcriptomic and chromatin dynamics of the human brain in PTSD. Nature 643, 744–754 (2025).

76. Mourtzi, N., Sertedaki, A. & Charmandari, E. Glucocorticoid Signaling and Epigenetic Alterations in Stress-Related Disorders. Int J Mol Sci 22 (2021).

77. Watkeys, O.J., Kremerskothen, K., Quide, Y., Fullerton, J.M. & Green, M.J. Glucocorticoid receptor gene (NR3C1) DNA methylation in association with trauma, psychopathology, transcript expression, or genotypic variation: A systematic review. Neurosci Biobehav Rev 95, 85–122 (2018).

78. McGowan, P.O. et al. Epigenetic regulation of the glucocorticoid receptor in human brain associates with childhood abuse. Nat Neurosci 12, 342–8 (2009).

79. Zhao, X. et al. PTSD and epigenetic aging: a longitudinal meta-analysis. Psychol Med 55, e142 (2025).

80. Wolf, E.J. et al. Traumatic stress and accelerated DNA methylation age: A meta-analysis. Psychoneuroendocrinology 92, 123–134 (2018).

81. Hannum, G. et al. Genome-wide methylation profiles reveal quantitative views of human aging rates. Mol Cell 49, 359–367 (2013).

